# Estimating SARS-CoV-2 seroprevalence and epidemiological parameters with uncertainty from serological surveys

**DOI:** 10.1101/2020.04.15.20067066

**Authors:** Daniel B. Larremore, Bailey K. Fosdick, Kate M. Bubar, Sam Zhang, Stephen M. Kissler, C. Jessica E. Metcalf, Caroline O. Buckee, Yonatan H. Grad

## Abstract

Establishing how many people have already been infected by SARS-CoV-2 is an urgent priority for controlling the COVID-19 pandemic. Patchy virological testing has hampered interpretation of confirmed case counts, and unknown rates of asymptomatic and mild infections make it challenging to develop evidence-based public health policies. Serological tests that identify past infection can be used to estimate cumulative incidence, but the relative accuracy and robustness of various sampling strategies has been unclear. Here, we used a flexible framework that integrates uncertainty from test characteristics, sample size, and heterogeneity in seroprevalence across tested subpopulations to compare estimates from sampling schemes. Using the same framework and making the assumption that serological positivity indicates immune protection, we propagated these estimates and uncertainty through dynamical models to assess the uncertainty in the epidemiological parameters needed to evaluate public health interventions. We examined the relative accuracy of convenience samples versus structured surveys to estimate population seroprevalence and found that sampling schemes informed by demographics and contact networks outperform uniform sampling. The framework can be adapted to optimize the design of serological surveys given particular test characteristics and capacity, population demography, sampling strategy, and modeling approach, and can be tailored to support decision-making around introducing or removing interventions.

## Introduction

Serological testing is a critical component of the response to COVID-19 as well as to future epidemics. Assessment of population seropositivity, a measure of the prevalence of individuals who have been infected in the past and developed antibodies to the virus, can address gaps in knowledge of the cumulative disease incidence. This is particularly important given inadequate viral diagnostic testing and incomplete understanding of the rates of mild and asymptomatic infections (*1*). In this context, serological surveillance has the potential to provide information about the true number of infections, allowing for robust estimates of case and infection fatality rates (*2*) and for the parameterization of epidemiological models to evaluate the possible impacts of specific interventions and thus guide public health decision-making.

The proportion of the population that has been infected by, and recovered from, the coronavirus causing COVID-19 will be a critical measure to inform policies on a population level, including when and how social distancing interventions can be relaxed. Individual serological testing may allow low-risk individuals to return to work, school, or university, contingent on the immune protection afforded by a measurable antibody response (*3,4*). At a population level, however, methods are urgently needed to design and interpret serological data based on testing of subpopulations, including convenience samples such as blood donors (*2, 5, 6*) and neonatal heel sticks, to reliably estimate population seroprevalence.

Three sources of uncertainty complicate efforts to learn population seroprevalence from subsampling. First, tests may have imperfect sensitivity and specificity. Estimates for COVID-19 tests on the market as of May 2020 reported specificity between 95% and 100% and sensitivity between 62% and 97% (Supplementary Table S1). Complicating this issue is the fact that sensitivity and specificity are, themselves, estimated from data (*7, 8*), which can lead to statistical confusion if uncertainty is not correctly propagated (*9*). Second, the population sampled will likely not be a representative random sample (*9*), especially in the first rounds of testing, when there is urgency to test using convenience samples and potentially limited serological testing capacity. Third, there is uncertainty inherent to any model-based forecast which uses the empirical estimation of seroprevalence, regardless of the quality of the test, in part because of the uncertain relationship between seropositivity and immunity (*10*).

A clear evidence-based guide to aid the design of serological studies is critical to policy makers and public health officials both for estimation of seroprevalence and for forward-looking modeling efforts, particularly if serological positivity reflects immune protection. To address this need, we developed a framework that can be used to design and interpret serological studies, with applicability to SARS-CoV-2. Starting with results from a serological survey of a given size and age stratification, the framework incorporates the test’s sensitivity and specificity and enables estimates of population seroprevalence that include uncertainty. These estimates can then be used in models of disease spread to calculate the effective reproductive number *R*_eff_, the transmission potential of SARS-CoV-2 under partial immunity, to forecast disease dynamics, and to assess the impact of candidate public health and clinical interventions. Similarly, starting with a pre-specified tolerance for uncertainty in seroprevalence estimates, the framework can be used to optimize the sample size and allocation needed. This framework can be used in conjunction with any model, including ODE models (*3, 11*), agent-based simulations (*12*), or network simulations (*13*), and can be used to estimate *R*_eff_ or to simulate transmission dynamics.

## Methods

### Design and modeling framework

We developed a framework for the design and analysis of serosurveys in conjunction with epidemiological models (Fig. 1), which can be used in two directions. In the forward direction, starting from serological data, one can estimate seroprevalence. While valuable on its own, seroprevalence can also be used as the input to an appropriate model to update forecasts or estimate the impacts of interventions. In the reverse direction, sample sizes can be calculated to yield seroprevalence estimates with a desired level of uncertainty and efficient sampling strategies can be developed based on prospective modeling tasks. The key methods include seroprevalence estimation, propagation of uncertainty through models, and model-informed sample size calculations.

**Figure 1:**
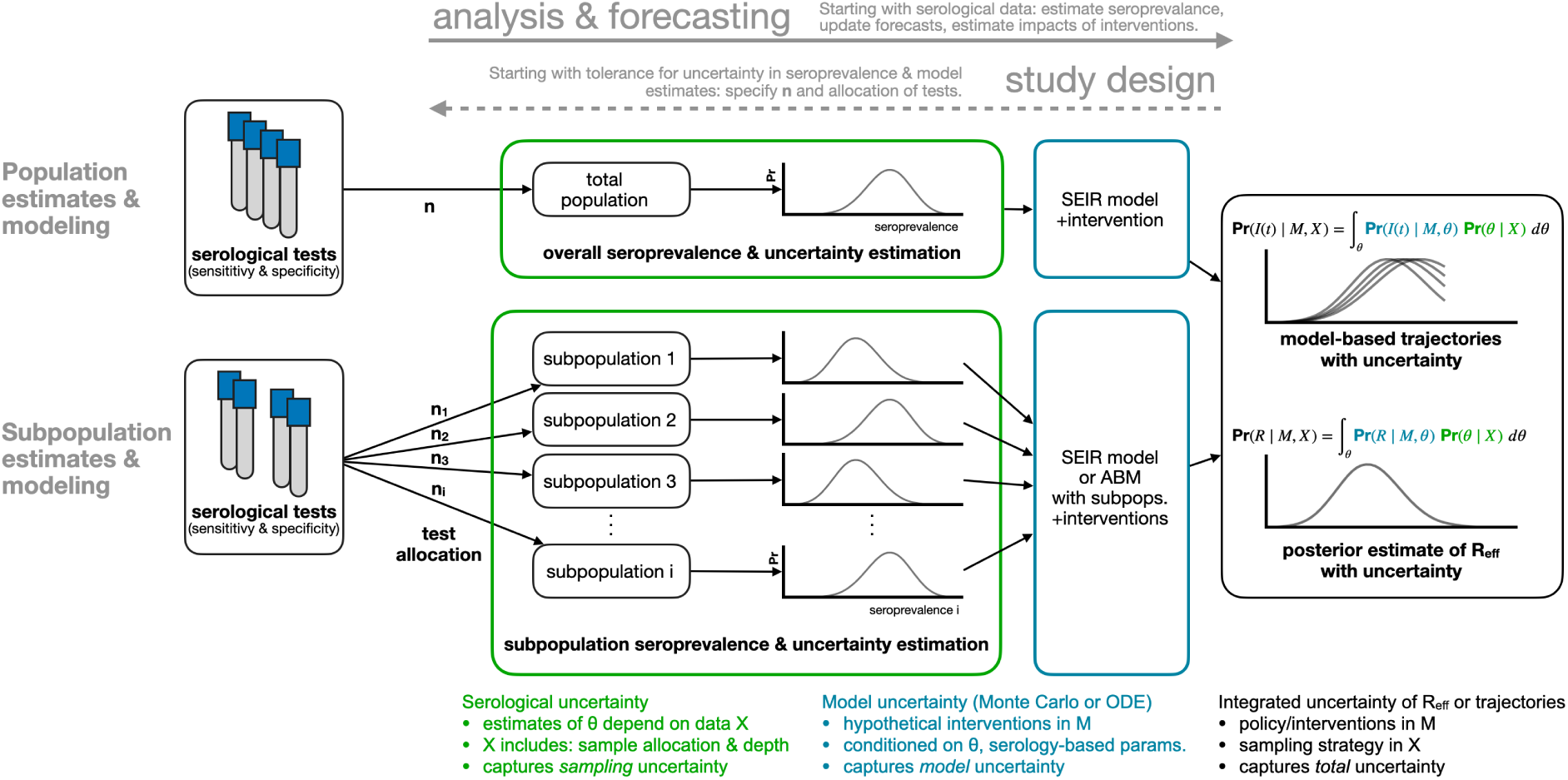
Framework for estimating seroprevalence and epidemiological parameters and the associated uncertainty, and for designing seroprevalence studies.

### Bayesian inference of seroprevalence

To integrate uncertainty arising from test sensitivity and specificity, we used a Bayesian model to produce a posterior distribution of seroprevalence that incorporates uncertainty associated with a finite sample size (Fig. 1; green annotations). We denote the posterior probability that the true population seroprevalence is equal to *θ*, given test outcome data *X* and test sensitivity and specificity characteristics, as Pr(*θ* | *X*). Because sample size and outcomes are included in *X*, and because test sensitivity and specificity are included in the calculations, this posterior distribution over *θ* appropriately handles uncertainty, and can be used to produce a point estimate of seroprevalence or a posterior credible interval. The model and sampling algorithm are fully described in Supplementary Text S1.

Sampling frameworks for seropositivity estimates are likely to be non-random and constrained to subpopulations. For example, convenience sampling (testing blood samples that were obtained for another purpose and are readily available) will often be the easiest and quickest data collection method (*14*). Two examples of such convenience samples are newborn heel stick dried blood spots, which contain maternal antibodies and thus reflect maternal exposure, and serum from blood donors (*2, 5, 6*). As a result, another source of statistical uncertainty comes from uneven sampling from a population.

To estimate seropositivity for all subpopulations based on a given sample (stratified, convenience, or otherwise), we specified a Bayesian hierarchical model that included a common prior distribution on subpopulation-specific seropositivities *θ*_*i*_ (Supplementary Text S1). In effect, this allowed seropositivity estimates from individual sub-populations to inform each other while still taking into account subpopulation specific testing outcomes. The joint posterior distribution of all subpopulation prevalences was sampled using Markov chain Monte Carlo (MCMC) methods (Supplementary Text S1). Those samples, representing posterior seroprevalence estimates for individual subpopulations, were then combined in a demographically weighted average to obtain estimates of overall seroprevalence, a process commonly known as poststratification (*8, 15*).

### Propagating serological uncertainty through models

In addition to estimating core epidemiological quantities (*16–18*) or mapping out patterns of outbreak risk (*19*), the posterior distribution of seroprevalence can be used as an input to any epidemiological model. Such models include the standard SEIR model, where the proportion seropositive may correspond to the recovered/immune compartment, as well as more complex frameworks such as an age-structured SEIR model incorporating interventions like school closures and social distancing (*20, 21*) (Fig. 1; blue annotations). We integrated and propagated uncertainty in the posterior estimates of seroprevalence and uncertainty in model dynamics or parameters using Monte Carlo sampling to produce a posterior distribution of epidemic trajectories or key epidemiological parameter estimates (Fig. 1; black annotations).

### Single-population SEIR model with social distancing and serology

To integrate inferred seroprevalence with uncertainty into a single-population SEIR model, we created an ensemble of SEIR model trajectories by repeatedly running simulations whose initial conditions were drawn from the seroprevalence posterior distribution. In particular, the seroprevalence posterior distribution was sampled, and each sample *θ* was used to inform the fraction of the population initially placed into the “recovered” compartment of the model. Thus, uncertainty in posterior seroprevalence was propagated through model outcomes, which were measured as epidemic peak timing and peak height. Social distancing was modeled by decreasing the contact rate between susceptible and infected model compartments. A full description of the model and its parameters can be found in Supplementary Text S2 and Supplementary Table S1.

### Age-structured SEIR model with serology

To integrate inferred seroprevalence with uncertainty into an age-structured SEIR model, we considered a model with 16-age-bins (0 − 4, 5 − 9, … 75 − 79). This model was parameterized using country-specific age-contact patterns (*22, 23*) and COVID-19 parameter estimates (*20*). The model, due to (*20*), includes age-specific clinical fractions and varying durations of preclinical, clinical, and subclinical infectiousness, as well as a decreased infectiousness for subclinical cases. A full description of the model and its parameters can be found in Supplementary Text S2 and Supplementary Table S1.

As in the single-population SEIR model, seroprevalence with uncertainty was integrated into the age-structured model by drawing samples from seroprevalence posterior to specify the fraction of each subpopulation placed into “recovered” compartments. Posterior samples were drawn from the age-stratified joint posterior distribution whose subpopulations matched the model’s subpopulations. For each set of posterior samples, the effective reproduction number *R*_eff_ was computed from the model’s next-generation matrix. Thus, we quantified both the impact of age-stratified seroprevalence (assumed to be protective) on *R*_eff_ as well as uncertainty in *R*_eff_.

### Serosurvey sample size and allocation for inference and modeling

The flexible framework described in Fig. 1 enables the calculation of sample sizes for different serological survey designs. To calculate the number of tests required to achieve a seroprevalence estimate with a specified tolerance for uncertainty, and to determine optimal test allocation across subpopulations in the context of studying a particular intervention, we treated the estimate uncertainty as a framework output and then sought to minimize it by improving the allocation of samples (Fig. 1, dashed arrow).

Uniform allocation of samples to subpopulations is not always optimal. It can be improved by i) increasing sampling in subpopulations with higher seroprevalence, and ii) increasing sampling in subpopulations with higher relative influence on the quantity to be estimated. This approach, which we term model and demographics informed (MDI), allocates samples to subpopulations in proportion to how much sampling them would decrease the posterior variance of estimates, i.e, 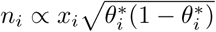 where 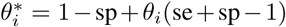 is the probability of a positive test in subpopulation *i* given test sensitivity (se), test specificity (sp), and subpopulation seroprevalence *θ*_*i*_, and *x*_*i*_ is the the relative importance of subpopulation *i* to the quantity to be estimated.

The sample allocation recommended by MDI varies depending on the information available and the quantity of interest. When the key quantity is overall seroprevalence, *x*_*i*_ is the fraction of the population in subpopulation When the key quantity is total infections *R*_eff_, or another quantity derived from compartmental models with subpopulations, *x*_*i*_ is the *i*th entry of the principal eigenvector of the model’s next generation matrix, including modeled interventions. If subpopulation prevalence estimates *θ*_*i*_ are unknown, sample allocation based solely on *x*_*i*_ is recommended. These methods are derived in Supplementary Text S3.

### Data Sources

Age distribution of U.S. blood donors was drawn from a study of Atlanta donors (*24*). Age distribution of U.S. mothers were drawn from the 2016 CDC Vital Statistics Report, using Massachusetts as a reference state (*25*). Daily age-structured contact data were drawn from Prem. et al (*23*). All data were represented using 5-year age bins, i.e. (0 − 4, 5 − 9,…,74 − 79). For datasets with bins wider than 5 years, counts were distributed evenly into the five-year bins. Serological test characteristics were collected from the websites of manufacturers and summarized in Supplementary Table S1. No attempt was made to test or validate manufacturer claims. Demographic data for the U.S. and India (analyzed in the manuscript) as well as other countries (provided in open-source code (*26*)) were downloaded from the 2019 United Nations World Populations Prospects report (*27*).

## Results

### Test sensitivity/specificity, sampling bias, and true seroprevalence influence the accuracy and robustness of estimates

We simulated serological data from a single population with seroprevalence rates ranging from 1% to 50% using the reported sensitivity (93%) and specificity (97.5%) of a test approved for sale in the EU (Supplementary Table S1), and with the number of samples ranging from 100 to 5000. We constructed Bayesian posterior estimates of seroprevalence, finding that, when seroprevalence is 10% or lower, around 1000 samples are necessary to es-timate seroprevalence to within two percentage points (Fig. 2). Tests with other characteristics required around 1000 tests (93.8% sensitivity, 95.6% specificity; Supplementary Fig. S1A) and 750 tests (97.2% sensitivity and 100% specificity; Supplementary Fig. S1B) to achieve the same uncertainty levels, relative to the minimum of around 650 tests for a theoretical test with perfect sensitivity and specificity (Supplementary Fig. S1C). In general estimates were most uncertain when true seropositivity was near 50%, the number of samples was low, and/or test sensitivity/specificity were low (Fig. 2 and Supplementary Fig. S1).

**Figure 2:**
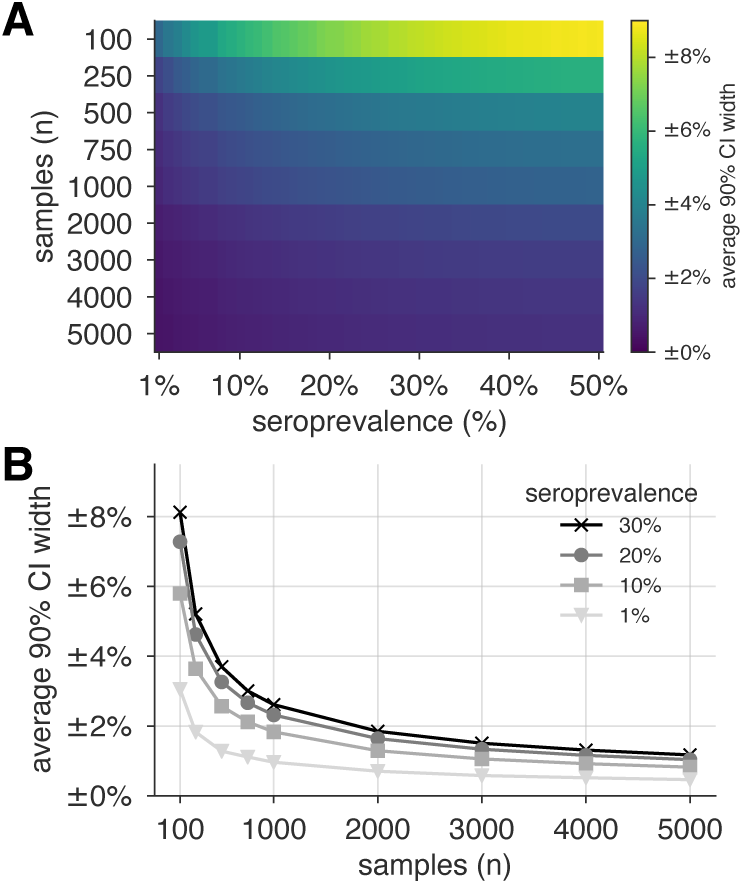
Uncertainty of population seroprevalence estimates as a function of number of samples and true population rate. Uncertainty, represented by the width of 90% credible intervals, is presented as ± seroprevalence percentage points in (A) a heatmap and (B) for selected seroprevalence values, based on a serological test with 93% sensitivity and 97.5% specificity (Supplementary Fig. S1 depicts results for other sensitivity and specificity values). 5000 samples are sufficient to estimate any seroprevalence to within a worst-case tolerance of ±1.3 percentage points, even with the imperfect test studied. Each point or pixel is averaged over 250 stochastic draws from the specified seroprevalence with the indicated sensitivity and specificity.

Next, we tested the ability of the Bayesian hierarchical model to infer both population and subpopulation seroprevalence. We simulated serological data from subpopulations for which samples were allocated and with heterogeneous seroprevalence levels (Supplementary Table S2). Test outcomes were randomly generated conditioning on the false positive and negative properties of the test being modeled (Supplementary Table S1). Test allocations across subpopulations were specified in proportion to age demographics of blood donations, deliver-ing mothers, uniformly across subpopulations, or according to an MDI allocation focused on minimized posterior uncertainty in *R*_eff_.

Credible intervals of the resulting overall seroprevalence estimates were influenced by the age demographics sampled, with the most uncertainty in the newborn dried blood spots sample set, due to the narrow age range for the mothers (Fig. 3). For such sampling strategies, which draw from only a subset of the population, our approach assumes that seroprevalence in each subpopulation does not dramatically vary and thus infers that seroprevalence in the unsampled bins is similar to that in the sampled bins but with increased uncertainty. Uncertainty was also influenced by the overall seroprevalence, such that the width of the 90% credible interval increased with higher seroprevalence for a given sample size. While test sensitivity and specificity also impacted uncertainty, central estimates of overall seropositivity were robust for sampling strategies that spanned the entire population. Note that the MDI sample allocation shown in Fig. 3 was optimized to estimate *R*_eff_, and thus, while it performs well, it is slightly outperformed by uniform sampling when used to estimate overall seroprevalence.

**Figure 3:**
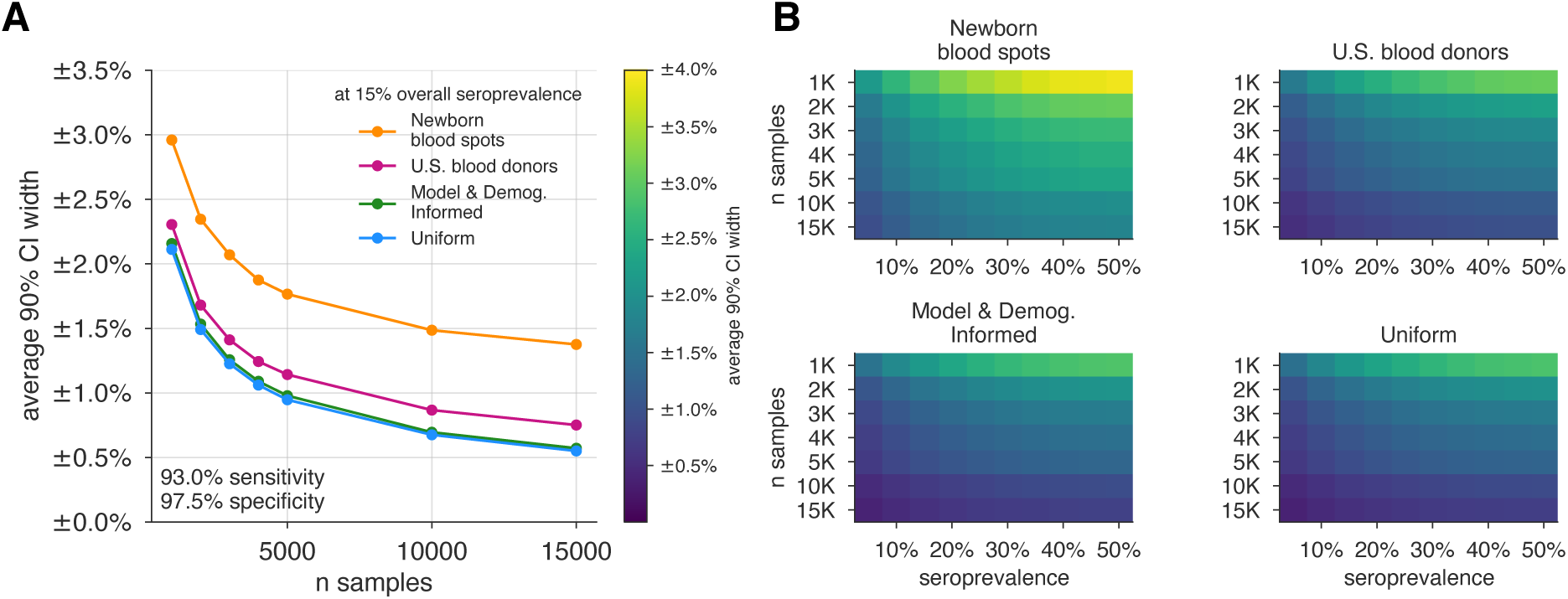
Uncertainty of overall seroprevalence estimates from convenience and formal sampling strategies. Uncertainty, represented by the width of 90% credible intervals, is presented as ± seroprevalence percentage points, based on a serological test with 93% sensitivity and 97.5% specificity (Supplementary Fig. S2 depicts results for other sensitivity and specificity values). (A) Curves show the decrease in average CI widths for 15% seroprevalence, illustrating the advantages of using uniform and MDI samples over convenience samples. (B) Heatmaps show average CI widths for various total sample counts and overall seroprevalence. Convenience samples derived from newborn blood spots or U.S. blood donors improve with additional sampling but retain baseline uncertainty due to demographics not covered by the convenience sample. For the estimation of overall seroprevalence, uniform sampling is marginally superior to this example of the model and demographic informed (MDI) sampling strategy, which was designed to optimize estimation of *R*_eff_. Each point or pixel is averaged over 250 stochastic draws from the specified seroprevalence with the indicated sensitivity and specificity.

### Seroprevalence estimates inform uncertainty in epidemic peak, timing, and reproductive number

Figure 4 illustrates how estimates of the height and timing of peak infections varied under two serological sampling scenarios and two hypothetical social distancing policies for a basic SEIR framework parameterized using seroprevalence data. Uncertainty in seroprevalence estimates propagated through SEIR model outputs in stages: larger sample sizes at a given seroprevalence resulted in a smaller credible interval for the seroprevalence estimate, which improved the precision of estimates of both the height and timing of the epidemic peak. Test characteristics also impacted model estimates, with more specific and sensitive tests leading to more precise estimates (Supplementary Fig. S3). Even estimations from a perfect test carried uncertainty corresponding to the size of the sample set (Supplementary Fig. S3).

**Figure 4:**
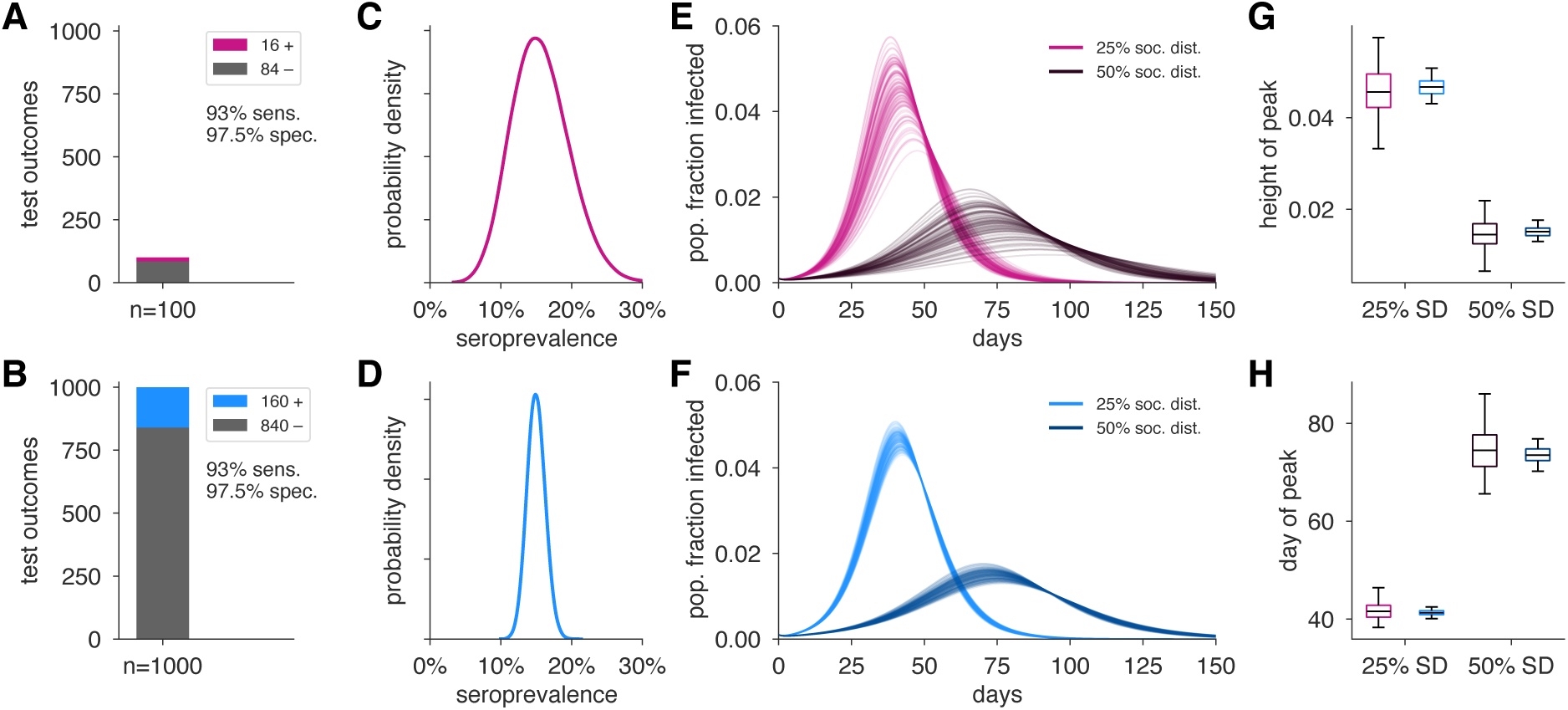
Uncertainty in serological data produces uncertainty in estimates of epidemic peak height and timing. Serological test outcomes for *n* = 100 tests (A; red) and *n* = 1000 tests (B; blue) produce (C,D) posterior seroprevalence estimates with quantified uncertainty. (E,F) Samples from the seroprevalence posterior produce a distribution of epidemic curves for scenarios of 25% and 50% social distancing (see Methods), leading to uncertainty in (G) epidemic peak and (H) timing which is mitigated in the *n* = 1000 sample scenario. Boxplot whiskers span 1.5×IQR, boxes span central quartile, lines indicate medians, and outliers were suppressed.

Figure 5 illustrates how the Bayesian hierarchical model extrapolates seroprevalence in sampled subpopulations, based on convenience samples from particular age groups or age-stratified serological surveys, to the overall population, with uncertainty propagated from these estimates to model-inferred epidemiological parameters of in-terest, such as *R*_eff_. Estimates from 1000 neonatal heel sticks or blood donations achieved more uncertain, but still reasonable, estimates of overall seroprevalence and *R*_eff_ as compared to uniform or demographically informed sample sets (Fig. 5). Here, convenience samples produced higher confidence estimates in the heavily sampled subpopulations, but high uncertainty estimates in unsampled populations through our Bayesian modeling framework. In all scenarios, our framework propagated uncertainty appropriately from serological inputs to estimates of overall seroprevalence (Fig 5I) or *R*_eff_ (Fig 5J). Improved test sensitivity and specificity correspondingly improved estimation and reduced the number of samples required (i) to achieve the same credible interval for a given seroprevalence and (ii) for estimates of *R*_eff_ (Supplementary Figs. S5 and S7).

**Figure 5:**
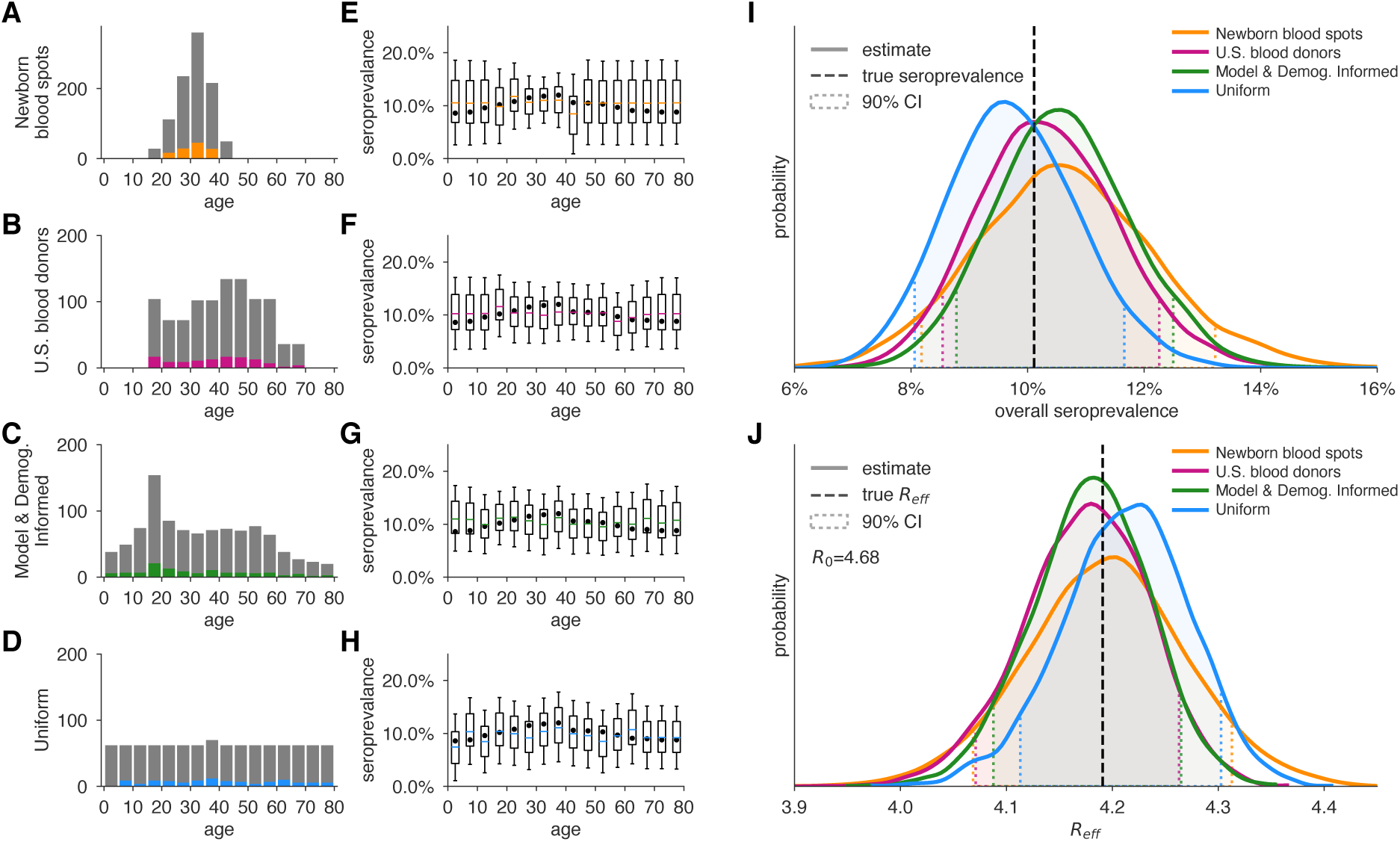
Convenience and formal samples provide serological and epidemiological parameter estimates. (A-D) For four sampling strategies, *n* = 1000 tests were allocated to age groups with negative tests (grey) and positive tests (colors) as shown, for a test with 93% sensitivity and 97.5% specificity. The MDI strategy shown was designed to optimize estimation of *R*_eff_. (E-H) Age-group seroprevalence estimates *θ*_*i*_ are shown as boxplots (boxes 90% CIs, whiskers 95% CIs); dots indicate the true values from which data were sampled. Note the decrease uncertainty for boxes with higher sampling rates. (I) Age-group seroprevalences were weighted by population demographics to produce overall seroprevalence estimates, shown as probability densities with 90% credible intervals shaded and highlighted with dashed lines. (J) Age-group seroprevalences were used to estimate *R*_eff_ under *status quo ante* contact patterns, shown as probability densities with 90% credible intervals shaded and highlighted with dashed lines. Dashed lines indicate true values from which the data were sampled. Each distribution depicts inference outcomes from a single set of stochastically sampled data; no averaging is done. Note that although uniform sample allocation produces a more confident estimate of overall seroprevalence, MDI produces a more confident estimate of *R*_eff_ since it allocates more samples to age groups most relevant to model dynamics.

If the subpopulations in the convenience sample have systematically different seroprevalence rates from the general population, increasing the sample size may bias estimates (Supplementary Figs. S4 and S7). This can be avoided using data from other sources or by updating the prior distributions in the Bayesian model with known or hypothesized relationships between seroprevalence of the sampled and unsampled populations.

### Strategic sample allocation improves estimates

We used the MDI strategy to design a study that optimizes estimation of *R*_eff_ and then tested the performance of the sample allocations against those resulting from blood donation and neonatal heel stick convenience sampling, as well as uniform sampling. As designed, MDI produced higher confidence posterior estimates (Fig. 5J, Supplementary Fig. S7). Importantly, because the relative importance of subpopulations in a model vary based on the hypothetical interventions being modeled (e.g., the re-opening of workplaces would place higher importance on the serological status of working-age adults), MDI sample allocation recommendations should be derived for multiple hypothetical interventions and then averaged to design a study from which the largest variety of high confidence results can be derived. To illustrate how such recommendations would work in practice, we computed MDI recommendations to optimize three scenarios for the contact patterns and demography of the U.S. and India, deriving a balanced sampling recommendation (Fig. 6).

**Figure 6:**
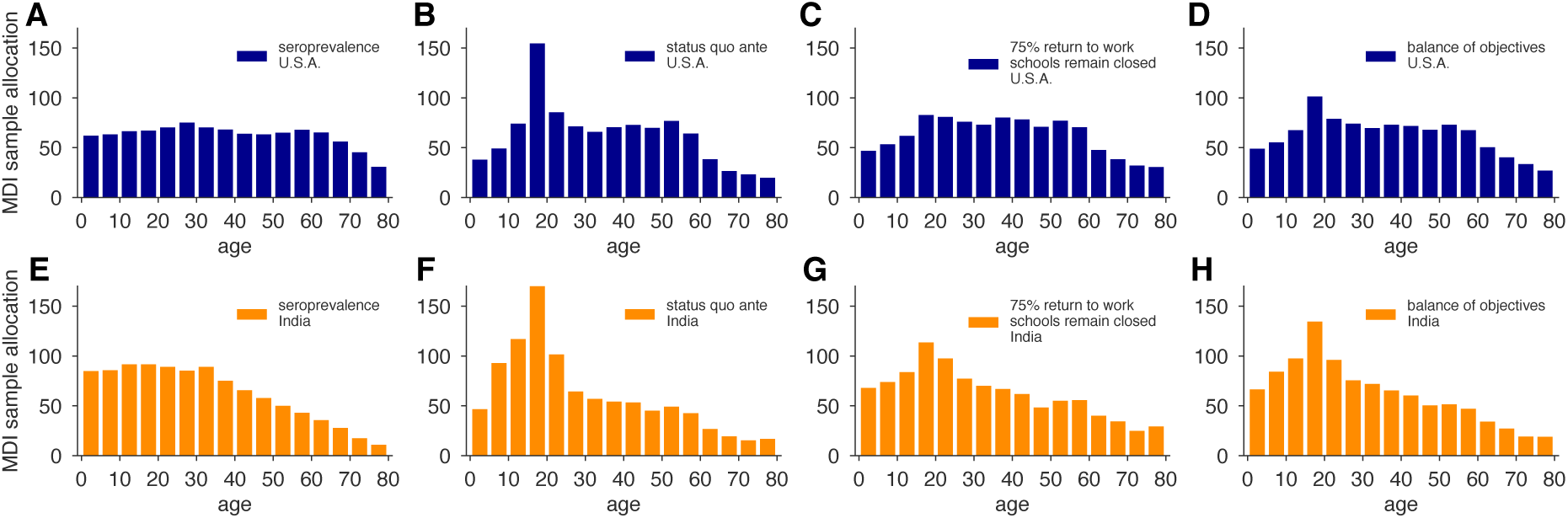
MDI sample allocations vary by demographics and modeling needs. Bar charts depict recommended sample allocation for three objectives, reducing posterior uncertainty for (A,E) estimates of overall seroprevalence, (B,F) predictions from an age-structured model with *status quo ante* contact patterns, (C,G) predictions from an age-structured model with modified contacts representing, relative to pre-crisis levels: a 20% increase in home contact rates, closed schools, a 25% decrease in work contacts and a 50% decrease of other contacts (*22, 23*), and (D,H) averaging the other three MDI recommendations to balance competing objectives. Data for both the U.S. (blue; A-D) and India (orange; E-H) illustrate the impact of demography and contact structure on strategic sample allocation. These sample allocation strategies assume no prior knowledge of subpopulation seroprevalences {*θ*_*i*_}.

## Discussion

There is a critical need for serological surveillance of SARS-CoV-2 to estimate cumulative incidence. Here, we presented a formal framework for doing so to aid in the design and interpretation of serological studies. We considered that sampling may be done in multiple ways, including efforts to approximate seroprevalence using convenience samples, as well as more complex and resource-intensive structured sampling schemes, and that these efforts may use one of any number of serological tests with distinct test characteristics. We incorporated into this framework an approach to propagating the estimates and associated uncertainty through mathematical models of disease transmission (focusing on scenarios where seroprevalence maps to immunity) to provide decision-makers with tools to evaluate the potential impact of interventions and thus guide policy development and implementation.

Our results suggest approaches to serological surveillance that can be adapted as needed based on pre-existing knowledge of disease prevalence and trajectory, availability of convenience samples, and the extent of resources that can be put towards structured survey design and implementation.

In the absence of baseline estimates of cumulative incidence, an initial serosurvey can provide a preliminary estimate (Fig. 2). Our framework updates the ‘rule of 3’ approach (*28*) by incorporating uncertainty in test characteristics and can further address uncertainty from biased sampling schemes (see Supplementary Text S4). As a result, convenience samples, such as newborn heel stick dried blood spots or samples from blood donors, can be used to estimate population seroprevalence. However, it is important to note that in the absence of reliable assess-ment of correlations in seroprevalence across age groups, extrapolations from these convenience samples may be misleading as sample size increases (Supplementary Figs. S4 and S6). Uniform or model and demographic informed samples, while more challenging logistically to implement, give the most reliable estimates. The results of a one-time study could be used to update the priors of our Bayesian hierarchical model and improve the inferences from convenience samples. In this context, we note that our framework naturally allows the integration of samples from multiple test kits and protocols, provided that their sensitivities and specificities can be estimated (*7,8*), which will become useful as serological assays improve in their specifications.

The results from serological surveys will be invaluable in projecting epidemic trajectories and understanding the impact of interventions. We have shown how the estimates from these serological surveys can be propagated into transmission models, incorporating model uncertainty as well. Conversely, to aid in rigorous assessment of particular interventions that meet accuracy and precision specifications, this framework can be used to determine the needed number and distribution of population samples via model and demographic informed sampling. Extensions could conceivably address other study planning questions, including sampling frequency (*29*).

There are a number of limitations to this approach that reflect uncertainties in the underlying assumptions of serological responses and the changes in mobility and interactions due to public health efforts (*30*). Serology reflects past infection, and the delay between infection and detectable immune response means that serological tests reflect a historical cumulative incidence (the date of sampling minus the delay between infection and detectable response). The possibility of heterogeneous immune responses to infection and unknown dynamics and duration of immune response mean that interpretation of serological survey results may not accurately capture cumulative incidence. For COVID-19, we do not yet understand the serological correlates of protection from infection, and as such projecting seroprevalence into models that assume seropositivity indicates immunity to reinfection may be an overestimate; models would need to be updated to include partial protection or return to susceptibility.

Use of model and demographic-informed sampling schemes are valuable for projections that evaluate interventions but are dependent on accurate parameterization. While in our examples we used POLYMOD and other contact matrices, these represent the *status quo ante*, and should be updated to the extent possible using other data, such as those obtainable from surveys (*22,23*) and mobility data from online platforms and mobile phones (*31–33*). Moreover, the framework could be extended to geographic heterogeneity as well as longitudinal sampling, if, for example, one wanted to compare whether the estimated quantities of interest (e.g., seroprevalence, *R*_eff_) differ across locations or time (*11, 19, 34*).

Overall, the framework here can be adapted to communities of varying size and resources seeking to monitor and respond to the SARS-CoV-2 pandemic. Further, while the analyses and discussion focused on addressing urgent needs, this is a generalizable framework that with appropriate modifications can be applicable to other infectious disease epidemics.

## Data Availability

All data and open-source code for this manuscript can be found in the github repository referenced in this paper.

https://github.com/LarremoreLab/covid_serological_sampling

## Acknowledgments

The authors wish to thank Nicholas Davies, Laurent H’bert-Dufresne, Johan Ugander, Arjun Seshadri, and the BioFrontiers Institute IT HPC group. The work was supported in part by the Morris-Singer Fund for the Center for Communicable Disease Dynamics at the Harvard T.H. Chan School of Public Health. Reproduction code is open source and provided by the authors (*26*).

## Supplementary Materials For

## S1 Bayesian inference of seroprevalence

### S1.1 Inference of seroprevalance in a sample using an imperfect test

If a serological test had perfect sensitivity and specificity, the probability of observing *n*_+_ seropositive results from *n* tests, given a true population seroprevalence *θ*, is given by the binomial distribution:

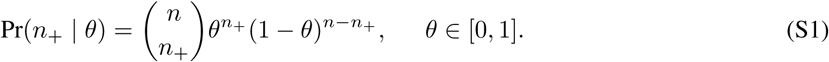

However, imperfect specificity and sensitivity require that we modify this formula. **For convenience, in the remainder of this Supplementary Text, we will use:**

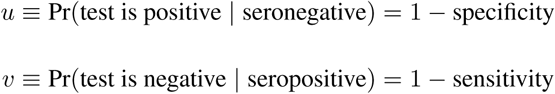

Using this notation, the probability that a single test returns a positive result, given *u, v*, and the true seroprevalence *θ*, is

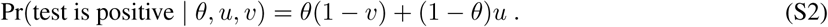

Substituting this per-sample probability into Eq. (S1) yields

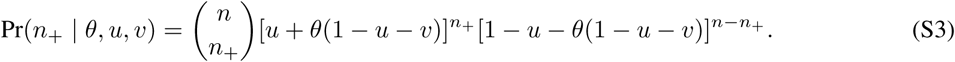

Finally, using Bayes’ Rule, we can write the posterior distribution over seropositivity *θ*, given the data, the test’s parameters (*35*), and an uninformative (uniform) prior on *θ*, yielding

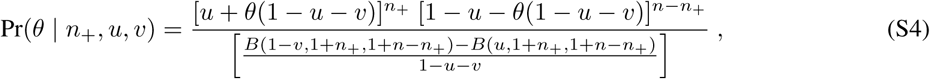

where *B* is an *incomplete beta function* without normalization. In practice, to sample from this distribution, one can use an accept-reject algorithm with, for example, a uniform proposal distribution and consider only the numerator of Eq. (S4). Alternatively, one can generate samples from a truncated Beta distribution using accept-reject sampling or an inverse cumulative distribution function method and these samples can be transformed to represent draws from Eq. (S4).

### S1.2 Bayesian estimation of seroprevalence across subpopulations

For a test with sensitivity 1 − *v* and specificity 1 − *u*, and given *n*_*i*+_ seropositive results from *n*_*i*_ tests in subpopulation *i*—set equal to zero for unsampled subpopulations—the posterior distribution over the vector of subpopulation seropositivities ***θ*** = {*θ*_*i*_} given all results ***n***_**+**_ = {*n*_*i*+_} is given by

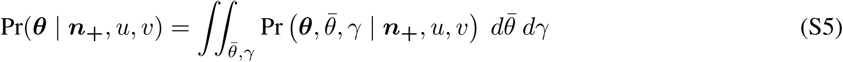

where we have included a hierarchy of priors. Specifically, the prior for each subpopulation seroprevalence was 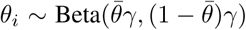, which has expectation 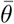 and variance 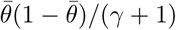. The hyperprior for the overall mean 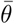 was uniform on the interval (0,1), allowing it to be dictated by the observed data. The hyperprior for the variance parameter was *γ* ∼ Gamma(*ν*, scale = *γ*_0_/*ν*), which has expected value *E*[*γ*] = *γ*_0_ and 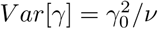.

### S1.3 Sampling from the Bayesian hierarchical model for subpopulation seroprevalences using MCMC

We sample from the joint posterior distribution inside the integral in Eq. (S5) using a Markov chain Monte Carlo (MCMC) algorithm, with univariate Metropolis-Hastings updates. We initialize the age-specific seroprevelance parameters at 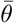 equal to the sample mean of the {*θ*_*i*_} and set *γ* = *γ*_0_. For each simulation, the MCMC algorithm was run for a total of 50, 100 iterations. The first 100 iterations were discarded and every 50th sample was saved to obtain 1, 000 samples from the joint posterior distribution. Code is open source and freely available (*26*). Diagnostic plots were produced and suggested these settings returned sufficient estimates of the posterior distribution.

## S2 Including protective seropositivity into models

### S2.1 Canonical SEIR model with social distancing and seropositivity

Let *S, E, I*, and *R* be the number of susceptible, exposed, infected, and recovered people in a population of size *N, S* + *E* + *I* + *R* = *N*. We model dynamics by

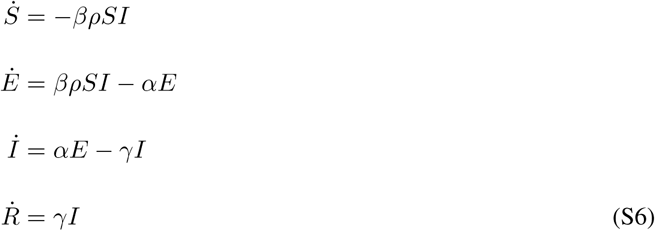

where *β, α*, and *γ* represent the rates of infection, symptom onset, and recovery, respectively, as in a standard SEIR model. To model social distancing we include the contact parameter *ρ* ∈ [0, 1] which modulates the fraction of social contacts between *S* and *I* populations that remain. Thus, *ρ* = 1 represents no social distancing while while *ρ* = 0.5 would represent a 50% reduction in contacts. In the simulations of this paper, only *ρ* = 0.5, 0.75 were considered as examples of dynamics.

To parameterize this model using seroprevalence, we made the modeling assumption that seropositive individuals are immune. Noting that this is only an assumption which at present requires in-depth research, we therefore placed seropositive individuals into the recovered group. In other words, for a seropositive fraction *θ*, with 10 individuals in the *E* and *I* compartments each, initial conditions would be,

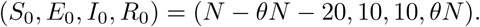

### S2.2 Canonical SEIR parameters and simulation details

Parameter values used in this study can be found in Supplementary Table S2. In prose, the model used transmission rate *β* = 1.75, exposure-to-infected rate *α* = 0.2, and recovery rate *γ* = 0.5, with no births or deaths, in a finite p opulation o f s ize *N* = 10, 0 00. S ocial d istancing w as i mplemented a s a c oefficient *ρ* = {0.5, 0.75}, corresponding to 50% and 25% social distancing, multiplying the contact rate between infected and susceptible populations. Integration was performed for 150 days with a timestep of 0.1 days. Initial conditions for (*S, E, I, R*) were (*N* − 20 − *θN*, 10, 10, *θN*), to simulate a fraction *θ* of recovered individuals, assumed to be immune. For each sampled value of *θ*, peak infection height and timing were extracted from forward-integrated timeseries.

### S2.3 Age-structured (POLYMOD) model with seropositivity

A model with 16-age-bins (0 − 4, 5 − 9, … 75 − 79) was parameterized using country-specific age-contact patterns (*22, 23*) and COVID-19 parameter estimates (*20*). The model includes age-specific c linical f ractions and varying durations of preclinical, clinical, and subclinical infectiousness, as well as a decreased infectiousness for subclinical cases (*20*).

Davies et al. define a next-generation matrix,

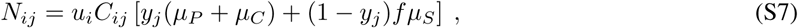

where *u*_*i*_ is the susceptibility of age group *i*; *C*_*ij*_ is the number of age-*j* individuals contacted by an age-*i* individual per day; *y*_*i*_ is the probability that an infection is clinical for an age-*i* individual; *µ*_*P*_, *µ*_*C*_, and *µ*_*S*_ are mean durations of preclinical, clinical, and subclinical infectiousness, respectively; and *f* is the relative infectiousness of subclinical cases (*20*). Values for all parameters are reported in Supplementary Table S2.

Protective seropositivity can be included in the model by multiplying *N*_*ij*_ as defined above by 1 − *θ*_*i*_, where *θ*_*I*_ is the seropositivity rate of age-group *i*. With this included term, we can modify Eq. (S7) as

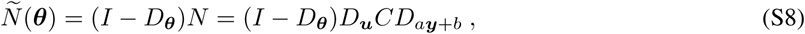

where *D*_***x***_ represents a diagonal matrix with entries *D*_*ii*_ = *x*_*i*_, and the constants are defined *a* = *µ*_*P*_ + *µ*_*C*_ − *fµ*_*S*_ and *b* = *µ*_*S*_.

The effective reproductive number is then the spectral radius *ρ* (i.e. the largest eigenvalue *λ*) of the next generation matrix:

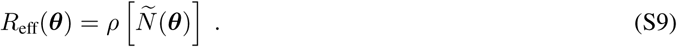

As written, Eq. (S9) represents a model component shown in Fig. 1 (blue annotations) as it maps parameters ***θ*** to a point estimate of *R*_eff_. As with the canonical SEIR model, uncertainty in the model parameters themselves can also be incorporated into overall uncertainty in *R*_eff_ via Monte Carlo.

### S2.4 Age-structured parameters and simulation details

Parameter values used in this study can be found in Supplementary Table S2, and were generally drawn from the work of Davies et al, and the sources therein. Published estimated contact matrices were used for India and the U.S. in the manuscript, with additional countries’ contact matrices shown in the accompanying open-source code (*26*).

## S3 Model and demographic informed (MDI) sampling

The calculations that follow rely on facts from optimization theory. We briefly review these here before applying these results in what follows.

Let ***n*** = (*n*_1_, …, *n*_*K*_). Suppose we want to minimize a function of the form

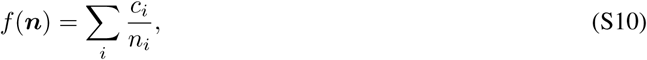

subject to the constraint that ∑_*i*_ *n*_*i*_ = *n*. Using the method of Lagrange multipliers, it can be shown that *f* (***n***) is minimized when 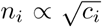. We apply this result below with various expressions for *c*_*i*_ to determine the optimal allocation of *n* tests across subpopulations in order to minimize the uncertainty of quantities of interest.

### S3.1 Minimizing posterior uncertainty for seroprevalence

Given age-specific seroprevalence estimates ***θ***, the estimate for overall seroprevalence is defined as *θ*_*pop*_ = ∑_*i*_ *d*_*i*_*θ*_*i*_, where *d*_*i*_ is the proportion of the population in group *i*. The uncertainty of this estimator depends on the uncertainties of the age-specific seroprevalences, which inherently depend on the number of tests *n*_*i*_ allotted to each subpopulation. Although the posterior uncertainties of the subpopulation seroprevalences are not available in closed form, we can nevertheless approximate them using the uncertainties in the corresponding maximum likelihood estimators. Here we consider the maximum likelihood estimators based on a separate binomial model for each subpopulation, i.e models of the form Eq. (S3) where *θ* is replaced by *θ*_*i*_. Note that this model assumes independence among the subpopulation seroprevalences.

The maximum likelihood estimate of *θ*_*i*_, given *n*_*i*,+_ positive tests out of *n*_*i*_ tests administered, is

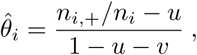

but this is only valid when both the numerator and denominator are positive, corresponding to a value of 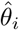 in the interval (0, 1). If the above estimator is computed and found to be negative, which happens when the fraction of tests that are positive is below the false positive rate, then the maximum likelihood lies at the endpoint, 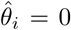. Similarly, if the estimator is found to be greater than one, 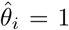. These estimators are undefined if no tests are allocated to group *i*, i.e. when *n*_*i*_ = 0.

Using the maximum likelihood estimators as proxies for the subpopulation posterior distributions, we can approximate the posterior variance of *θ*_*pop*_ as

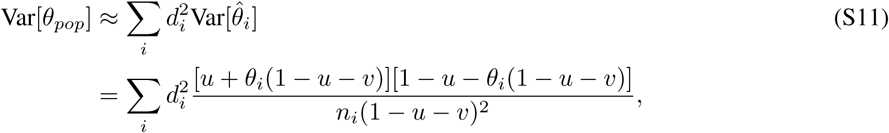

where *θ*_*i*_ is the true seroprevalence of group *i*. This variance equation has the form of Eq. (S10) and thus the optimal allocation of samples is given by

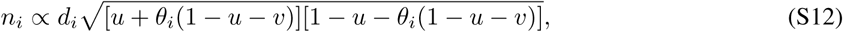

where multiplicative constants have been absorbed into the proportion. In the absence of knowledge about the true subpopulation seroprevalences ***θ***, we recommend simply allocating samples with respect to the demographic information: *n*_*i*_ ∝ *d*_*i*_.

### S3.2 Minimizing posterior uncertainty for modeling

When the primary quantity of interest is the output from a model, improved test allocation strategies can be developed by leveraging the model structure. For example, suppose the goal is accurate estimation of the total number of infected individuals at some future time point *t*. To avoid confusion with the identity matrix *I* or the subpopulation index *i*, let Let 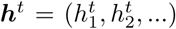 denote the vector containing the number of infected individuals within each subpopulation and let the total number of infected individuals be 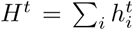. Using the next generation matrix defined in Eq. (S7) and modification as in Eq. (S8), the next generation matrix updates the vector of infected individuals per subpopulation as

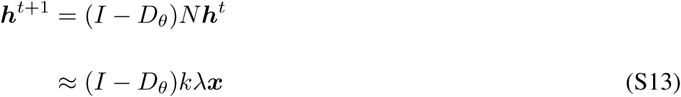

where ***x*** represents the eigenvector of *N* corresponding to the largest eigenvalue *λ*, and *k* is a scalar *k* = ***x***^*T*^ ***h***^*t*. 1^ There are two helpful interpretations of this equation. First, the vector ***x*** is the principal “direction” of the next generation matrix, and repeated iterations of the dynamics in a large population will result in infected fractions that are proportional to ***x***. In the above, we approximate the effect of *N* on ***h*** as *kλ****x***, an approximation which is better when *λ* is well separated from the second eigenvalue *λ*_2_. Measurements of *λ*/|*λ*_2_| for models considered in this manuscript ranged from 2 to 4.

A second interpretation of this result appeals to the notion of the next generation matrix *N* as a *network* in which the nodes are infected subpopulations and the directed links *N*_*ij*_ explain the effects of an infection at node *j* on future infections at node *i*. In this network dynamical system, by calculating ***x*** we have computed the *eigenvector centralities* of the network’s nodes (*36*), which are a measure of the importance of each subpopulation in the network.

With these preliminary calculations in mind, we turn to the estimation of *H*^*t*^. Because 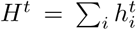, and because the values 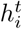 are all functions of a random variable ***θ***, *H*^*t*^ is also a random variable. Our goal is to minimize its variance by strategically allocating finite samples in order to minimize the *important* posterior variances among the elements of ***θ***. In plain language, some of the subpopulations are more important in shaping future disease dynamics than others, so MDI will preferentially allocate more samples to those subpopulations in a principled manner, which we now derive.

As in Eq. (S11), we approximate the posterior variance of ***θ*** by the posterior variance of the corresponding maximum likelihood estimator 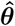. This results in the following approximation of the variance of the total number infected:

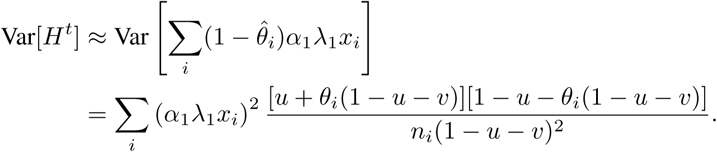

where *x*_*i*_ is the *i*th element of the principal eigenvector ***x***. The first expression is obtained by using the approximation in Eq. (S13). The resulting variance expression has the form of Eq. (S10) and thus, ignoring constants, the optimal allocation of samples is given by

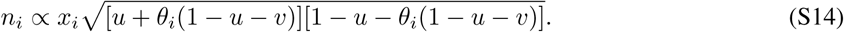

In the absence of knowledge about the true subpopulation seroprevalences ***θ***, we recommend simply allocating samples with respect to the entries of the principal eigenvector: *n*_*i*_ ∝ *x*_*i*_.

## S4 Impact of sensitivity and specificity on the “Rule of 3”

Suppose we have a perfect test (*u* = *v* = 0) and when we perform *n* tests, zero are positive. The maximum likelihood estimate of the seroprevalence would be 0. (*28*) proposed a simple upper 95% confidence bound on true seroprevalence equal to 3/*n*.

The derivation of this rule is motivated by the following question: “What is the maximum seroprevalence under which the probability of observing zero positives in *n* tests is less than or equal to 5%?”. Briefly, the probability of a negative test is *θ* and thus the probability of observing *n* negative tests is (1 − *θ*)^*n*^. Setting this equal to 0.05 and solving for *θ*, we find *θ* = 1 − .05^1/*n*^ ≈ 3/*n*, where the approximation is based on the power series representation of the exponential function.

Now, let’s consider what happens if sensitivity and specificity are not equal to one and again zero positive tests are observed. The probability of a negative test is then 1 − *u* − *θ*(1 − *u* − *v*). An upper 95% confidence bound on the true seroprevalence is then

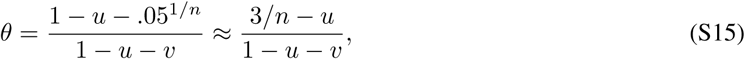

where the approximation is derived in a similar manner. Notice if *u* > 3/*n*, this upper bound is less than zero. This occurs when there is inconsistency between the specified false positive rate *u* and the observed data; namely, this occurs when *n* is large enough that we would have expected at least one false positive.

Even if seroprevalence is zero, we expect to observe some number positive tests simply due to imperfect test specificity. Suppose we observe *n*_+_ positive tests from a sample of *n*. An approximate upper 95% confidence bound on the true seroprevalence:

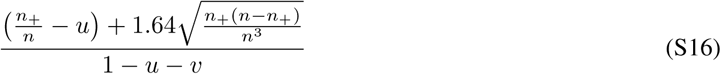

## Supplementary Figures

**Figure S1:**
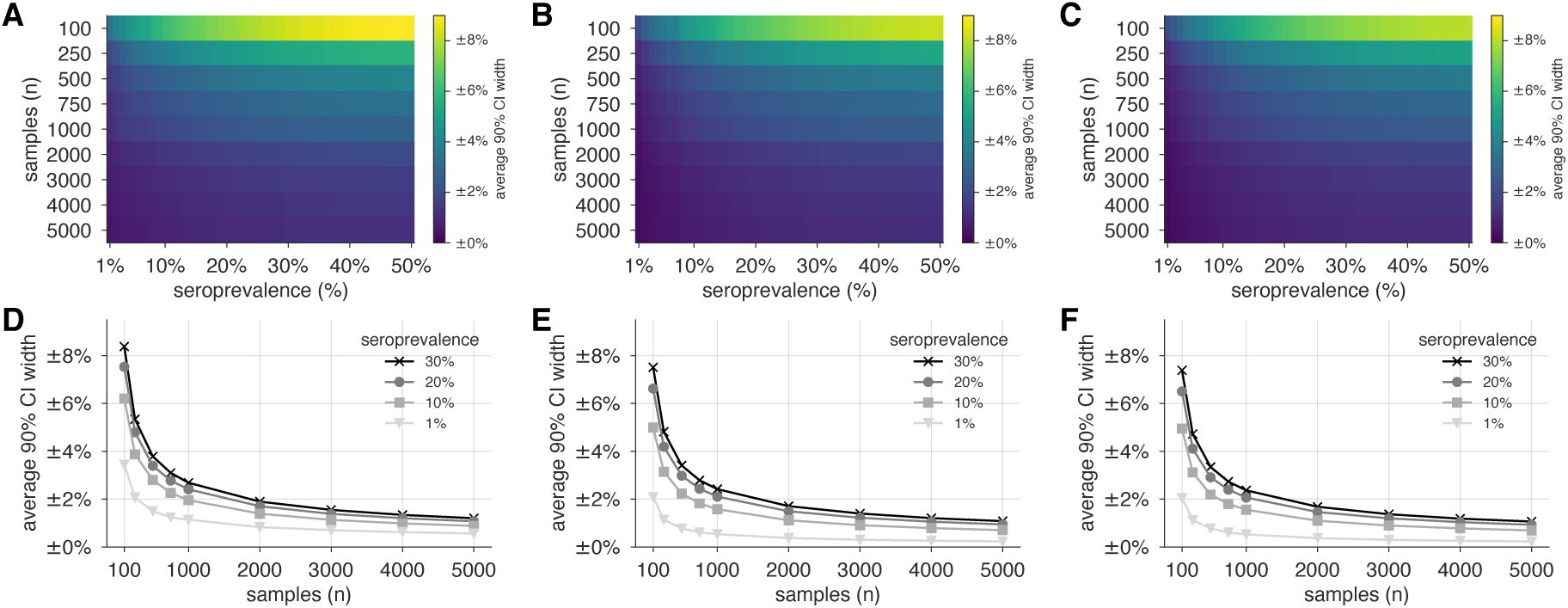
Uncertainty of population seroprevalence estimates as a function of number of samples and true population rate. Uncertainty, represented by the width of 90% credible intervals, is presented as ± seroprevalence percentage points in heatmaps and for selected seroprevalence values, based on a serological tests with (A,D) 93.8% sensitivity and 95.6% specificity, matching the claims of a Cellex test, (B,E) 97.2% sensitivity and 100% specificity, matching the claims of an Aytu IgG test, (C,F) 100% sensitivity and specificity, representing an ideal test, complementing the results for a test with 93% sensitivity and 97.5% specificity shown in the main text (Fig. 2). See Supplementary Table S1 for details on serological test kits.

**Figure S2:**
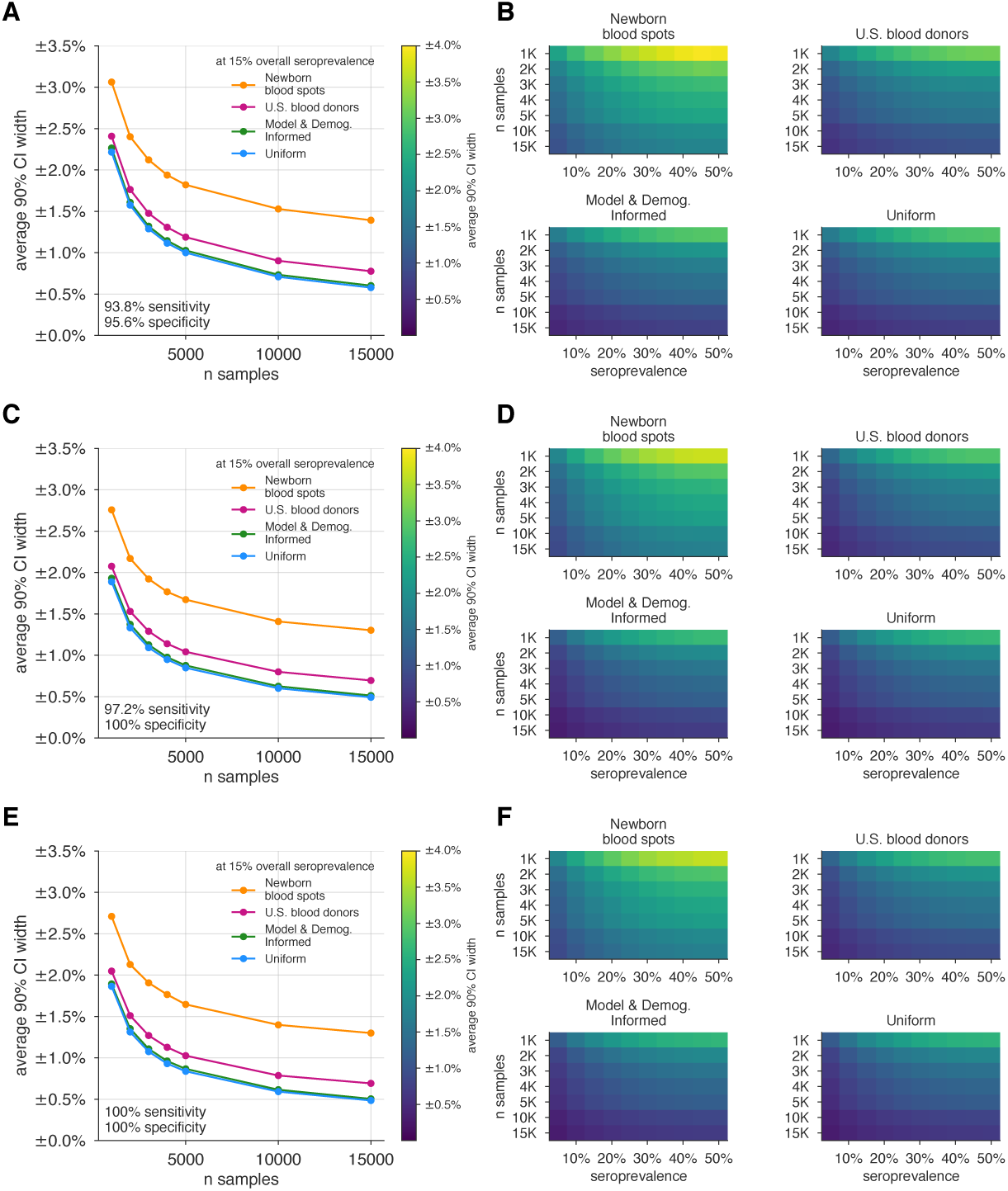
Uncertainty of overall seroprevalence estimates from convenience and formal sampling strategies. Uncertainty, represented by the width of 90% credible intervals, is presented as ± seroprevalence percentage points, based on a serological tests with (A,B) 93.8% sensitivity and 95.6% specificity, matching the claims of a Cellex test, (C,D) 97.2% sensitivity and 100% specificity, matching the claims of an Aytu IgG test, (E,F) 100% sensitivity and specificity, representing an ideal test. complementing the results for a test with 93% sensitivity and 97.5% specificity shown in the main text (Fig. 3). (A,C,E) Curves show the decrease in average CI widths for 15% seroprevalence, illustrating the advantages of using uniform and MDI samples over convenience samples. (B,D,F) Heatmaps show average CI widths for various total sample counts and overall seroprevalence. Convenience samples derived from newborn blood spots or U.S. blood donors improve with additional sampling but retain baseline uncertainty due to demographics not covered by the convenience sample. For the estimation of overall seroprevalence, uniform sampling is marginally superior to this example of the model and demographic informed (MDI) sampling strategy, which was designed to optimize estimation of *R*_eff_. Each point or pixel is averaged over 250 stochastic draws from the specified seroprevalence with the indicated sensitivity and specificity.

**Figure S3:**
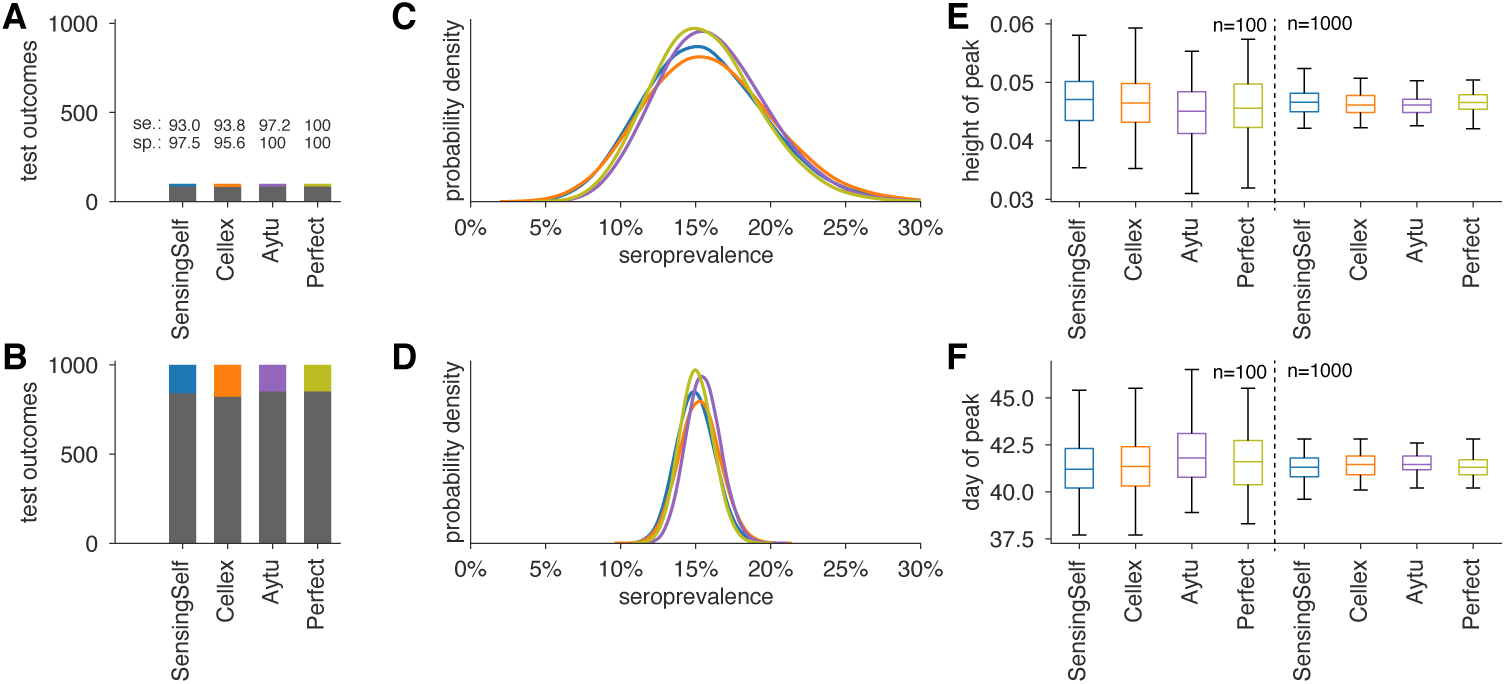
Uncertainty in serological data produces uncertainty in estimates of epidemic peak height and timing, even when the test has perfect sensitivity and specificity. Serological test outcomes for (A) *n* = 100 tests and (B) *n* = 1000 tests produce are shown as bar graphs for four tests with sensitivity and specificity values as indicated. Serological test samples were not generated stochastically but instead according to expectation to highlight how sensitivity and specificity affect inference. Posterior seroprevalence estimates for (C) *n* = 100 and (D) *n* = 1000 scenarios reveal that Bayesian estimate place posteriors over the correct values (15%) but with uncertainty that depends on *n* (compare C to D) and on test characteristics (compare peak heights of yellow and purple to blue and orange). Samples from the seroprevalence posterior produce a distribution of epidemic curves for scenarios of 25% and 50% social distancing (see Methods), leading to uncertainty in (E) height of epidemic peak and (F) timing of epidemic peak. Uncertainty is mitigated but not eliminated in the *n* = 1000 scenario, just as uncertainty is mitigated but not eliminated using a perfect serological test. Boxplots reflect 100 samples from SEIR dynamimcs; whiskers span 1.5×IQR, boxes span central quartile, lines indicate medians, and outliers not shown. See Methods for SEIR simulation details and parameters.

**Figure S4:**
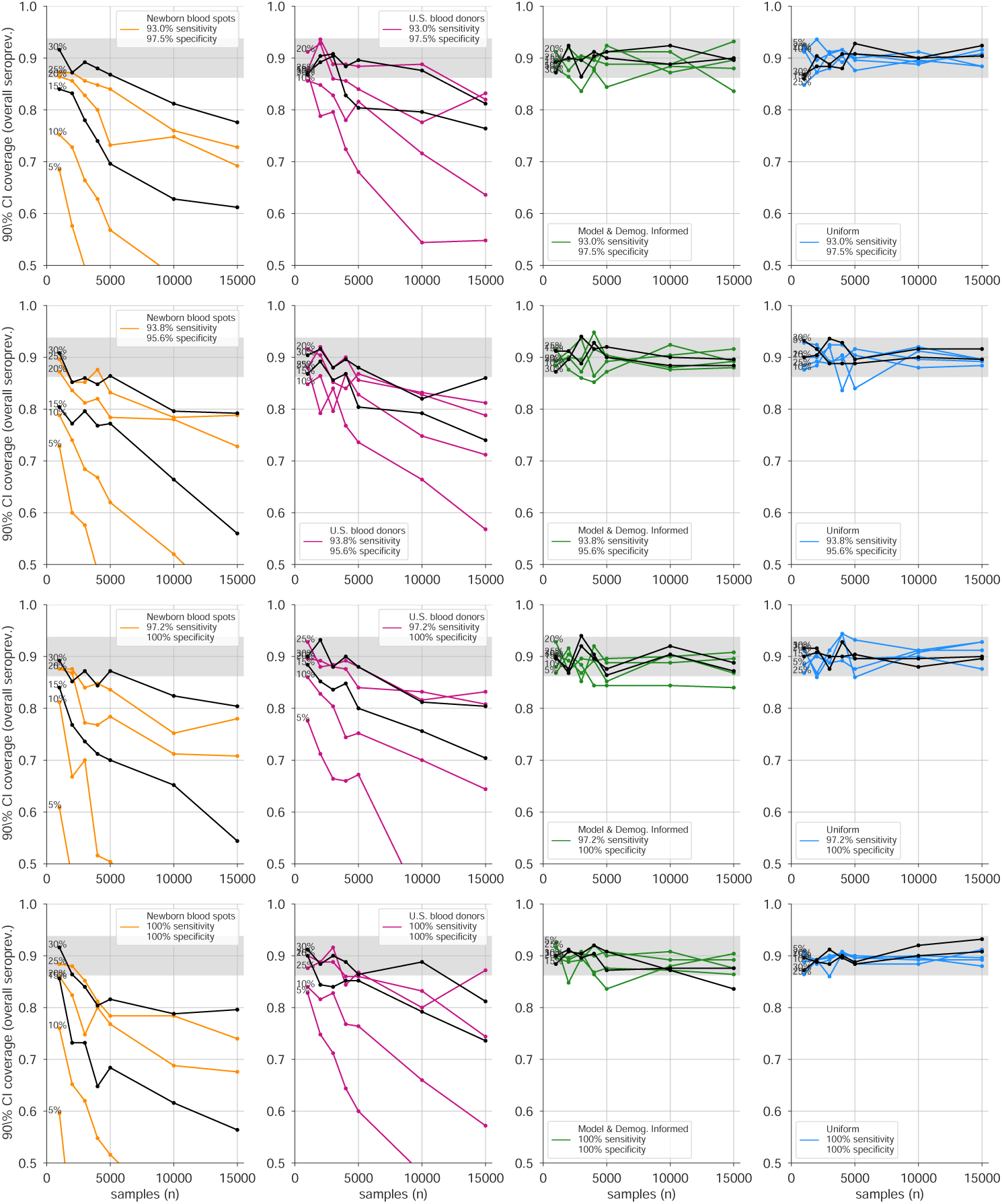
Credible interval coverage for overall seroprevalence estimates using four sampling strategies and four serological test kits. Credible interval coverage, defined as the fraction of posterior credible intervals that covered the true parameter used to generate the data, are shown for four sampling strategies (columns, colors) and four test kits (rows), with sensitivity and specificity values as indicated; see legends. Each point represents the fraction of credible intervals which covered the planted value for the indicated overall seroprevalence value (see annotations on plots) at the specified number of serological samples *n*, out of a total of 250 independent trials. The estimated coverage from a perfectly calibrated posterior will have coverage fractions within 0.9 ± 0.37 (grey bands) 95% of the time. Some seroprevalence values are plotted in black simply to guide the eye. The MDI strategy shown was designed to optimize estimation of *R*_eff_.

**Figure S5:**
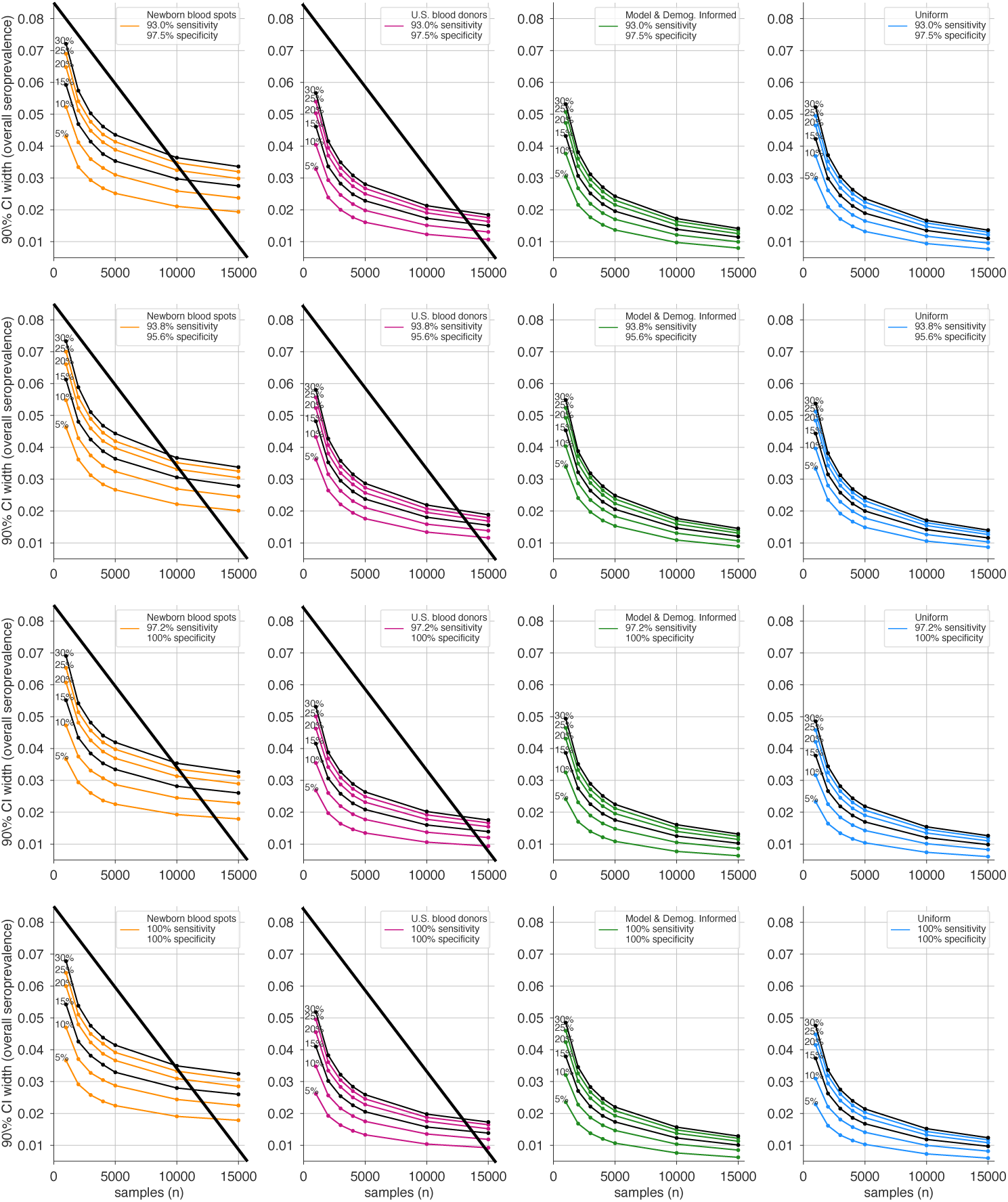
Average credible interval width for overall seroprevalence estimates using four sampling strategies and four serological test kits. Credible intervals were calculated for data generated according to four sampling strategies (columns, colors) and four test kits (rows), with sensitivity and specificity values as indicated; see legends. Each point represents the average width of the intervals for the indicated overall seroprevalence value (see annotations on plots) at the specified number of serological samples *n*, out of a total of 250 independent trials. Some seroprevalence values are plotted in black simply to guide the eye. The MDI strategy shown was designed to optimize estimation of *R*_eff_. Sampling strategies that resulted in posterior credible intervals with inaccurate coverage (see Supplementary Fig. S4) are crossed out.

**Figure S6:**
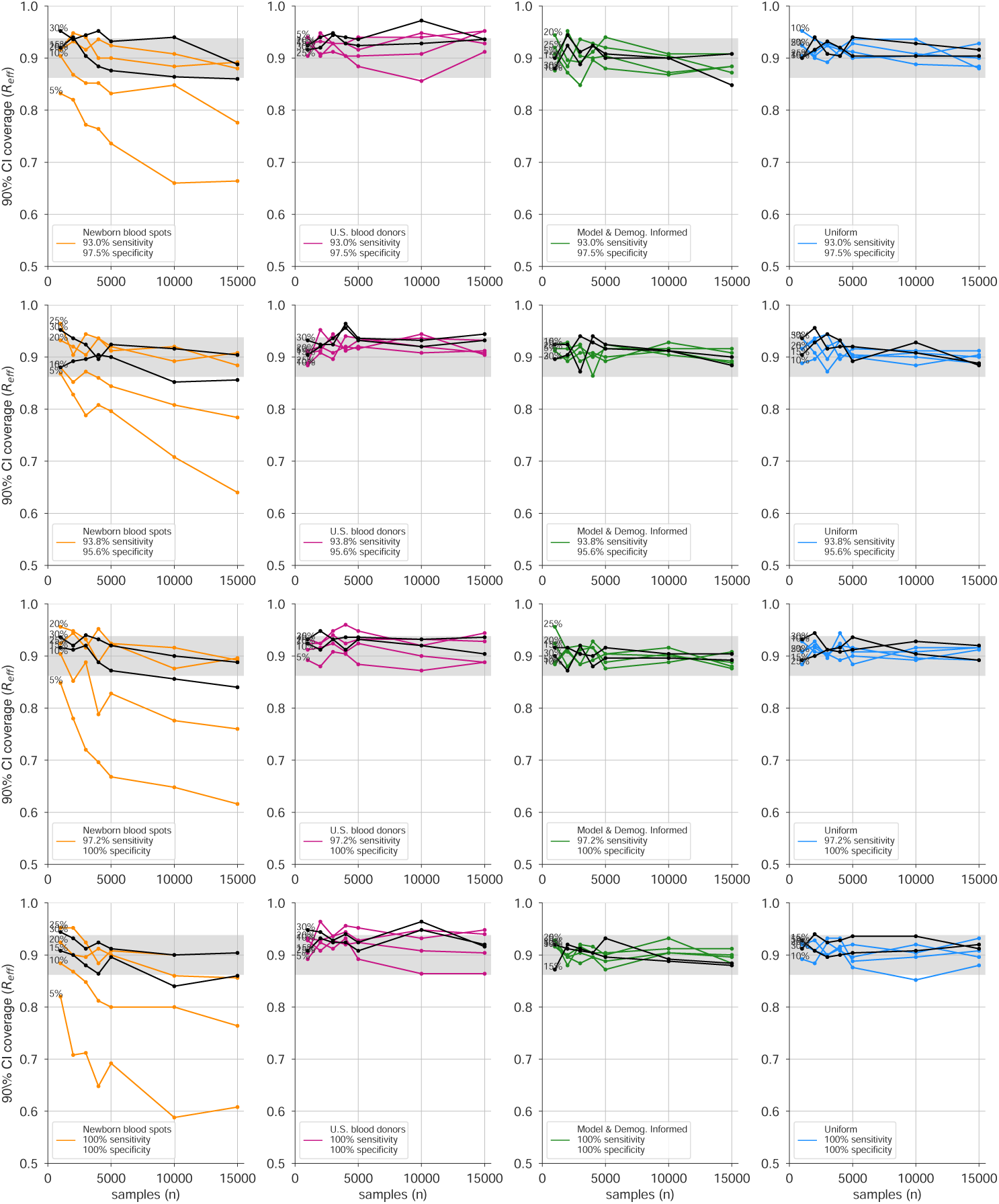
Credible interval coverage for *R*_eff_ estimates using four sampling strategies and four serological test kits. Credible interval coverage, defined as the fraction of posterior credible intervals that covered the true parameter used to generate the data, are shown for four sampling strategies (columns, colors) and four test kits (rows), with sensitivity and specificity values as indicated; see legends. Each point represents the fraction of credible intervals which covered the planted value for the indicated overall seroprevalence value (see annotations on plots) at the specified number of serological samples *n*, out of a total of 250 independent trials. The estimated coverage from a perfectly calibrated posterior will have coverage fractions within 0.9 ± 0.37 (grey bands) 95% of the time. Some seroprevalence values are plotted in black simply to guide the eye. The MDI strategy shown was designed to optimize estimation of *R*_eff_.

**Figure S7:**
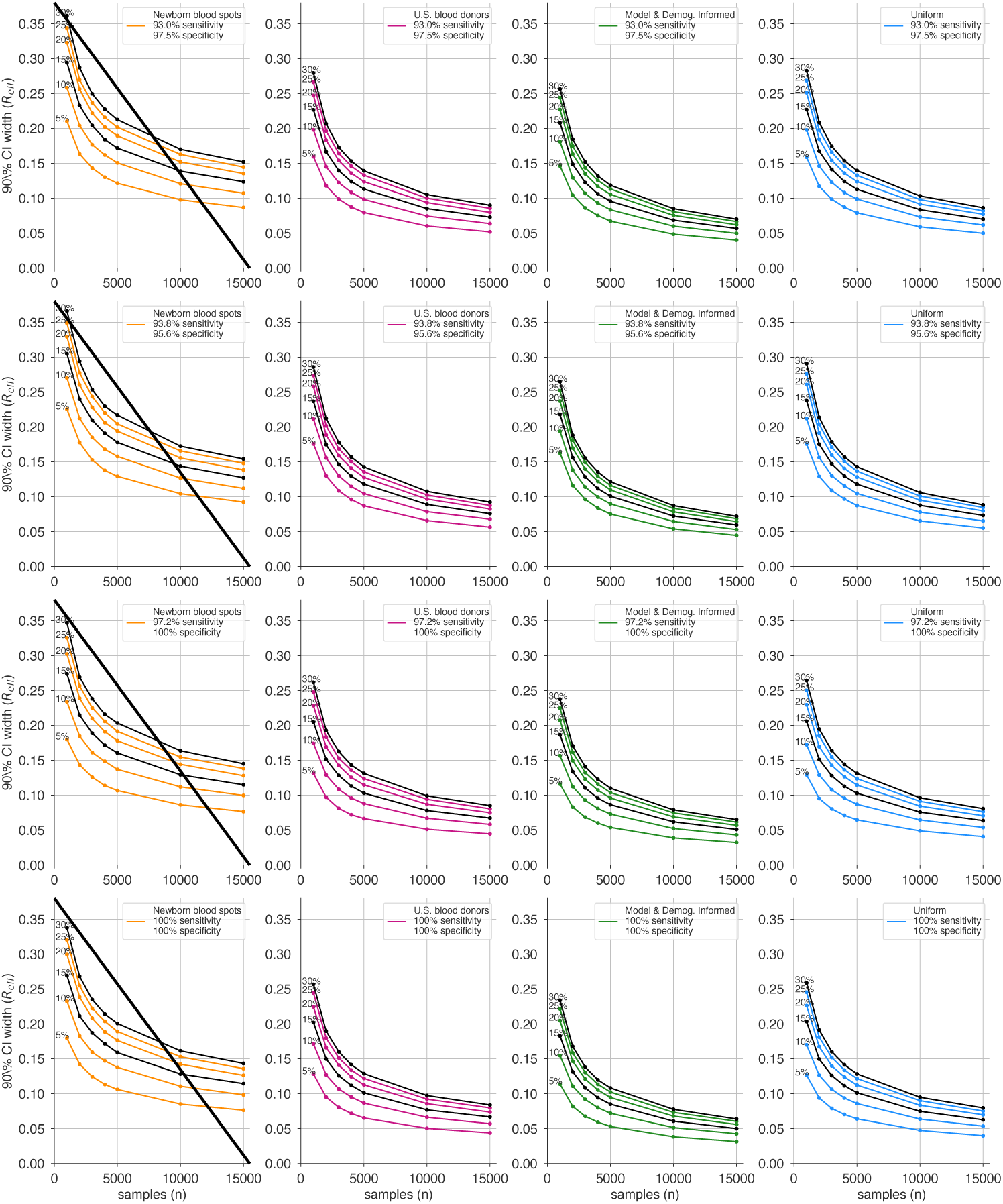
Average credible interval width for *R*_eff_ estimates using four sampling strategies and four serological test kits. Credible intervals were calculated for data generated according to four sampling strategies (columns, colors) and four test kits (rows), with sensitivity and specificity values as indicated; see legends. Each point represents the average width of the intervals for the indicated overall seroprevalence value (see annotations on plots) at the specified number of serological samples *n*, out of a total of 250 independent trials. Some seroprevalence values are plotted in black simply to guide the eye. The MDI strategy shown was designed to optimize estimation of *R*_eff_. Sampling strategies that resulted in posterior credible intervals with inaccurate coverage (see Supplementary Fig. S6) are crossed out.

## Supplementary Tables

**Table S1:** Serological tests used in this study. Sensitivity and specificity values were taken from manufacturer’s claims as of April 9, 2020, compiled by the Johns Hopkins Center for Health Security^1^.

| Manufacturer | Type of Test | Sens. | Spec. | Sales Approval |
| --- | --- | --- | --- | --- |
| *Sensing Self | IgG | 93 | 97.5 | CE |
| Sensing Self | IgM | 82 | 96 | CE |
| †Cellex Inc. | IgM & IgG | 93.8 | 95.6 | FDA, CE |
| †Aytu Biosciences/Orient Gene Biotech | IgG | 97.2 | 100 | CE, China |
| Aytu Biosciences/Orient Gene Biotech | IgM | 87.9 | 100 | CE, China |
| ScanWell Health/INNOVITA | IgM & IgG | 87.3 | 100 | China |
| SD Biosensor | IgM & IgG | 82 | 97 | Korea |
| Liming Bio | IgG | 82 | 100 | CE, IVD |
| Liming Bio | IgM | 62 | 100 | CE, IVD |
| Shenzhen Yhlo Biotech Company | IgG | 90 | 95 | CE, IVD |
| Shenzhen Yhlo Biotech Company | IgM | 95 | 95 | CE, IVD |

\* included only in main text modeling, calculations, and figures.
† included in Supplementary modeling, calculations, and figures.
<sup>1</sup> http://www.centerforhealthsecurity.org/resources/COVID-19/serology/Serology-based-tests-for-COVID-19.html

**Table S2:** Parameter values used in models. This table is divided into two sections. The top section corresponds to the parameters of the single-population SEIR model. The bottom section corresponds to the parameters used in the age-structured SEIR model. Contact matrices *C*_*ij*_ used in this manuscript were, in particular, those corresponding to the United States of America and India. Values for *y*_*i*_, the probability that an infection is clinical for an age-*i* individual, were generated by using three control points for young, middle and old age, then interpolating between them with a cosine-smoothing function, as described in (*20*). Equations for models can be found in Supplementary Text. Test kit sensitivity and specificity values are provided in Supplementary Table S1.

| Parameter | Meaning | Value | Reference |
| --- | --- | --- | --- |
| $\alpha$ | transition rate $E \rightarrow I$ | 0.2 | |
| $\beta$ | infectiousness | 1.75 | |
| $\gamma$ | recovery rate | 0.5 | |
| $\rho$ | social distancing effects | 0.5, 0.75 | |
| $\theta$ | seropositive fraction | various | see Methods |
| $u_i$ | Susceptibility of age group $i$ | 0.078 | (20) |
| $y_i$ | Probability that an infection is clinical for an age-$i$ individual | [0.056, 0.056, 0.056, 0.064, 0.100, 0.162, 0.240, 0.322, 0.398, 0.455, 0.486, 0.535, 0.723, 0.740, 0.740, 0.740] | (20)<br>(see caption) |
| $C_{ij}$ | Number of age-$j$ individuals contacted by an age-$i$ individual per day | various | (22)<br>(see caption) |
| $\mu_p$ | Mean duration of preclinical infectiousness | 2.4 days | (20, 37) |
| $\mu_c$ | Mean duration of clinical infectiousness | 3.2 days | (20, 38) |
| $\mu_s$ | Mean duration of subclinical infectiousness | 7 days | (20) |
| $f$ | Relative infectiousness of subclinical cases | 50% | (20) |
| $\theta_i$ | Seropositivity of age-$i$ subpopulation | $\tilde{\theta} + [-0.014, -0.012, -0.004, 0.002, 0.008, 0.015, 0.018, 0.020, 0.006, 0.005, 0.003, -0.003, -0.009, -0.010, -0.012, -0.012]$, with various $\tilde{\theta}$, representing the average seroprevalence | |

## Footnotes

1 The next generation matrix *N* is non-negative and satisfies the conditions of the Perron-Frobenius theorem which means that it has a largest eigenvalue *λ*—for a next generation matrix, *R*_0_ = *λ*—which is greater than or equal to all other eigenvalues, with a corresponding eigenvector *x* of non-negative components. This means that repeated applications of *N* to any initial vector that is not orthogonal to *x* will become increasingly parallel to *x* at a rate of *λ*/ *λ*_2_ per iteration, where *λ*_2_ is the second largest eigenvalue of *N*. This is the basis of the so-called Power Method which repeatedly applies the matrix to find the largest eigenvalue and its corresponding eigenvector.

